# Individualized, self-supervised deep learning for blood glucose prediction

**DOI:** 10.1101/2023.08.19.23294318

**Authors:** Johannes Fuest, Marco Tacke, Leander Ullman, Peter Washington

## Abstract

The current standard for monitoring blood glucose levels in diabetes patients are continuous glucose monitoring (CGM) devices, which are costly and carry the risk of complications, such as allergic reactions or skin irritations from the adhesive used to attach the CGM sensor to the skin. CGM devices are also highly visible and can thus act as a discomforting disease-marker for diabetes patients. To mitigate these issues, we develop and test a novel method that is able to predict blood glucose levels with only non-invasive predictor variables and a very small number of target variable measurements by using individualization and self-supervised deep learning. Using only a single blood glucose measurements per week, our method (6387.47 glucose-specific MSE) outperforms traditional deep learning performed with hourly measurements (8191.23 glucose-specific MSE). Across eight experiments where blood glucose measurements are more than one hour apart, our approach outperforms traditional deep learning without exception. Our findings suggest that self-supervised, individualized deep learning could provide an avenue towards alternatives to CGM devices that would be less costly, non-invasive, and thus more accessible.

## Introduction

Diabetes Mellitus is a metabolic disease characterized by chronic hyperglycemia due to the body’s inability to either adequately produce or efficiently utilize insulin [1]. Symptoms of the condition can include mortality, blindness, kidney failure and an overall decreased quality of life [2]. Today, there are over 400 million diabetes patients worldwide, with a further 350 million at risk of diabetes [3]. One in five adults over the age of 65 suffer from diabetes globally [4]. In the US, the condition disproportionately affects ethnic minorities and low-income populations [5]. Outside the US, low- and middle-income countries health systems are exposed to increased financial strain due to rising diabetes treatment costs [6]. Changes in lifestyles, urbanization, and an aging population will continue to exacerbate this crisis, with patient numbers expected to rise beyond 600 million by 2045 [7], making diabetes one of the most pressing global health challenges in need of innovative, low-cost solutions.

There is no cure for diabetes, with patients instead reliant on blood glucose monitoring, insulin injections and other measures for treatment [8]. Continuous glucose management (CGM) is a technique that measures a patient’s glucose levels in short intervals, in order to inform insulin dosages and other treatments. It offers more detailed glucose information than traditional self-monitoring of blood glucose (SMBG) [9]. While CGM has established itself as one of the most effective glucose monitoring techniques [10], one disadvantage is that it requires long-term implanted sensors, which risk infection or other adverse reactions [11], whilst also carrying significant up-front costs. As a consequence, CGM devices have not been widely adopted in many parts of the world [12] and are unsuitable in some settings, such as intensive care units [13].

To bypass the downsides of CGM devices, we propose a non-invasive method to predict blood glucose levels based on individualized, self-supervised deep learning that requires only a low number of blood glucose measurements, such as a single measurement per day or week, and no expensive hardware to perform well. We show that when blood glucose data is only available in large intervals, such as on a weekly basis, our approach, which combines the advantages of self-supervised deep learning and individualization, strongly outperforms traditional supervised deep-learning. This ability to perform well when glucose labels are scarce, is highly relevant, as it opens up the possibility of replacing CGM devices entirely with models trained using methods similar to our approach. Widespread use of techniques based on this idea could lead to cost savings, infection prevention, and more widespread access to diabetic care, potentially improving care for millions of patients.

## Related Work

Theree are many past examples of machine learning applications in diabetes, covering areas such as diagnosis [14] [15], as well as treatment methods [16] [17]. This study falls into the category of work attempting to use machine learning for continuous glucose monitoring, which is a key part of diabetes treatment. There have been other efforts to apply personalized machine learning to this problem. Alexiou at al. (2021) [18] proposed a personalized approach using three types of regression trees. However, this study included neither deep learning methodologies, nor self-supervised learning. There have been other studies using deep learning to predict blood glucose levels, but these involved either multisensor data [19] [20] or invasive data collected using expensive, state-of-the-art medical equipment [21], whereas we only use data from non-invasive sources. Li et al. (2020) [22] use similar input variables to this study, but use a different architecture based on recurrent convolutional neural networks and do not include a self-supervised component. Other studies have applied self-supervised learning to various medical topics [23], but to our knowledge we are the first to apply self-supervised learning to blood glucose prediction. Our study thus makes a new contribution to the field of machine learning for blood glucose prediction, by using only non-invasive, cheaply collected measurement data for prediction, and relying on self-supervised learning to enhance performance when label data is scarce.

Beyond the direct context of glucose prediction, this study relates to previous work on developing deep learning techniques for time series data, as well as applying self-supervised learning to time series problems. It builds on the framework proposed by Zhao et al. (2017) [24] for time series predictions using convolutional neural networks and demonstrates its suitability to continuous glucose monitoring. There have been many studies that successfully employ this approach to problem domains, such as energy consumption [25] and algorithmic trading [26]. Other studies have also used self-supervised and transfer learning in a time series setting with a similar architecture in areas including flood prediction [27].

This work further adds to a growing body of research into personalized machine learning within medicine. A wide range of medical conditions and diseases, as well as overall health monitoring systems have the potential to be revolutionized by big-data and machine learning over the coming years [28]. Increasingly granular measurements of various health aspects through wearable devices, and continuously improving cloud infrastructure are making diagnoses and predictions more precise and scalable. While personalization efforts in many areas of medicine are still in their infancy in terms of clinical adoption [29], there is a broad consensus about their vast potential [30]. Within diabetes care, there have been investigations of the potential for personalized medicine applications in diagnosis [31], treatment regimen [32], long-term average glucose levels [33], as well as genetic risk factors [34]. This study complements these approaches, by applying an advanced machine learning technique on patient-specific data labels, resulting in a personalized model for every patient.

## Materials and methods

### Data Source

The datasets used in this project are sourced from Jaeb et al. [35]. The authors of the dataset aimed to investigate whether an automated insulin management system could be used to safely manage diabetic patient’s blood glucose levels from home. The dataset is publicly available and contains blood glucose related data collected during two clinical phases of the study. The dataset comes in the form of 30 text files with different structures generated by different systems (the insuline pump, the glucose measurement device, a centralized computer, and the facilitators of the study). The study involved 30 participants, of which 14 completed the entirety of the two trials performed by the authors. The features we use for our analysis are measured blood glucose levels, which are our target variable, as well insulin amounts injected, exercise times, meal sizes, and liquid glucose consumed orally. All of these data points include timestamps. Blood glucose measurements of the patients were taken in roughly five-minute intervals.

The primary challenge associated with the data is the inconsistent and irregular intervals at which the different variables are reported. Time intervals between data points are inconsistent and there are temporal gaps in the data. We solve this by setting the regular glucose measurements as the level of observation and aggregating all other predictors into five-minute intervals for the 24-hour window *prior* to each glucose measurement. This results in 288 five-minute intervals for each of our four predictors. Thus, for every glucose measurement in the original study, we have 288 preceding five-minute windows of meal size, liquid glucose consumption, insulin injected, and exercise as predictors, with each 5-minute window describing the sum of the predictor variables at that time. Our final dataset after this preprocessing contains 845,696 rows, each with a timestamp, a glucose measurement, the 4 * 288 predictor columns described above, as well as a patient identifier.

We note that there is a large amount of redundancy in the dataset, because each row represents a 24-hour window that is shifted by five minutes in comparison with the preceding row, with the exception of instances where blood glucose labels are more than 5 minutes apart due to gaps in the data. We also note here that the CGM device used to generate the blood glucose labels for our analysis are not exact measurements, but should be considered estimates of current blood glucose levels. Furthermore, we note that only 14 of 30 patients in the original study participated in both clinical phases. Since the phases differed in length, in behavior of the patients, in the data generation techniques, as well as in the settings of the glucose pumps, we find that the data quality and consistency in phase two is far superior to that in phase one. Because of these complications, our final data preprocessing step was to exclude the 16 patients who participated only in phase one.

### Model Architecture

To evaluate how both individualization and self-supervised learning affect our performance for blood glucose level predictions, we test different training approaches which are outlined in the next subsection, using the same architecture across experiments to ensure comparability. Our model architecture is visualized in Figure 1. The model first takes in the four 288-interval predictors and feeds them into separate 1D convolutional layers, each including batch normalization and dropout. The output of these layers is then flattened and concatenated before being fed through a series of fully connected layers until a single scalar value is output representing the blood glucose prediction the model makes.

**Fig 1.**
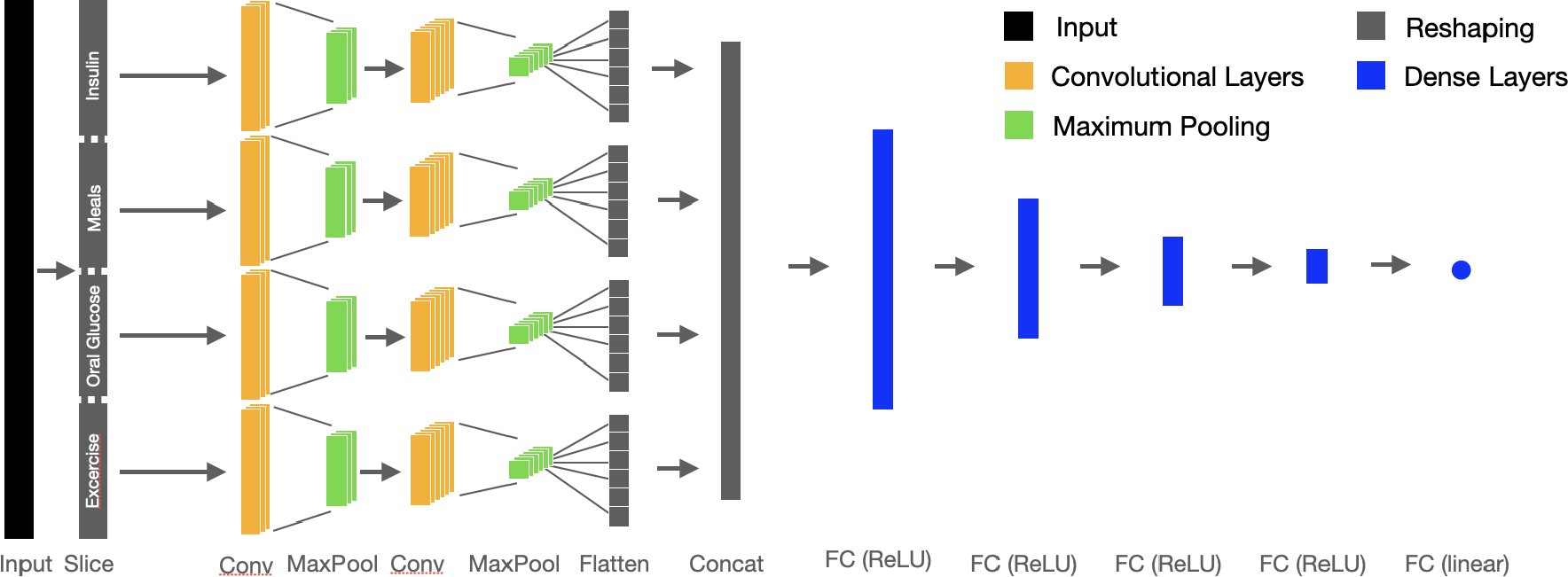
Model Architecture for All Experiments: We first feed the four time series predictors through a series of two 1D convolutional layers with batch normalization and dropout. We then concatenate the output from this and use linear layers to arrive at a single output.

This architecture builds on the framework proposed by Zhao et al. (2017) [24] for time series prediction using convolutional neural networks. We consider this a state-of-the art approach for time series prediction and do not include other architecture types in our analysis, as our discussion centres around training approaches rather than comparing varying model architectures or optimizing a particular architecture. Full details of our model structure, including layer dimensions, are provided in the appendix in Figure 1.

### Training Approach

Our novel training approach is centered around combining two main ideas. First, we hypothesize that building one model per patient, rather than training a model on all patients and hoping that it generalizes to other patients’ idiosyncrasies, improves performance. Individualization has yielded positive results in many other precision medicine tasks, and could thus also be beneficial in glucose monitoring [36]. Secondly, we hypothesize that employing self-supervised learning for our use case has the potential to further enhance performance. Self-supervised learning means first training the model to learn a task related to but distinct from the downstream task, based only on the predictor data, and then using the model parameters resulting from this training stage as the starting point for the supervised portion of training. The intuition behind this is that the self-supervised learning stage allows the model to learn about features of the system in question, which in this case is the individual patient. This later helps the model to perform better on the downstream task, which in our case is blood glucose prediction. Rather than measuring blood glucose every five minutes through an implanted sensor, we envision a future in which blood glucose only needs to be measured on a daily or even weekly basis. In such a setting with very few labels, we hypothesize that model performance on blood glucose prediction can be significantly enhanced by starting model training with a self-supervised training phase on the more abundant, non-invasive predictor data.

The degree to which self-supervised learning enhances performance is heavily dependent on the features the model learns during self-supervised training [37]. Due to the sparsity of the predictors in our dataset, the self-supervised task we use is to predict the sum of each of the four predictor variables over the next two hours given a row of our dataset. We initially considered a more traditional approach whereby each row of the predictors is used to predict the next row, but early results showed that this approach worsened performance. Across exercise, insulin, and liquid glucose consumption, the average value in our dataset is 0.062, and over 90% of all entries for mealsize are 0.0, since patients only recorded meal intakes in a handful of five minute intervals over any given 24 hour period. Because of this, we found that learning to predict the next row of our sparse dataset reduced a high number of weights of the model close to 0, which worsened performance on the downstream task, where labels range between 50 and 400. Pivoting to sums of our four predictors over the next 24 entries (corresponding to the next two hours) solved this problem. When training for this task, the output dimension of the last layer of the model is adjusted to output four values instead of the single output in the downstream task of glucose prediction. Self-supervised learning techniques similar to this have been successfully applied in many other domains [38].

In order to successfully combine the advantages of self-supervised learning and individualization in our setting, there is a trade-off to consider. Self-supervised learning works best when predictor data is abundant. Combining it directly with individualization may be problematic, since using predictor data from only one out of 14 patients in our dataset greatly reduces the amount of predictor data available. This harms self-supervised learning, as it is directly dependent on the abundance of the predictor data. For our proposed approach, we thus perform self-supervised learning on all patients’ data in the first phase of training. On completion, we then use the parameters from this training phase as a starting point for a traditional phase of supervised deep learning on all patients’ blood glucose labels. Finally, we leverage the advantages of individualization in the third training phase, by running supervised learning using only the individual patients’ data, which fine-tunes the model to the individual patient’s idiosyncrasies. The details of our training approach are summarized in figure 2 together with the details of baselines approaches we compare to.

**Fig 2.**
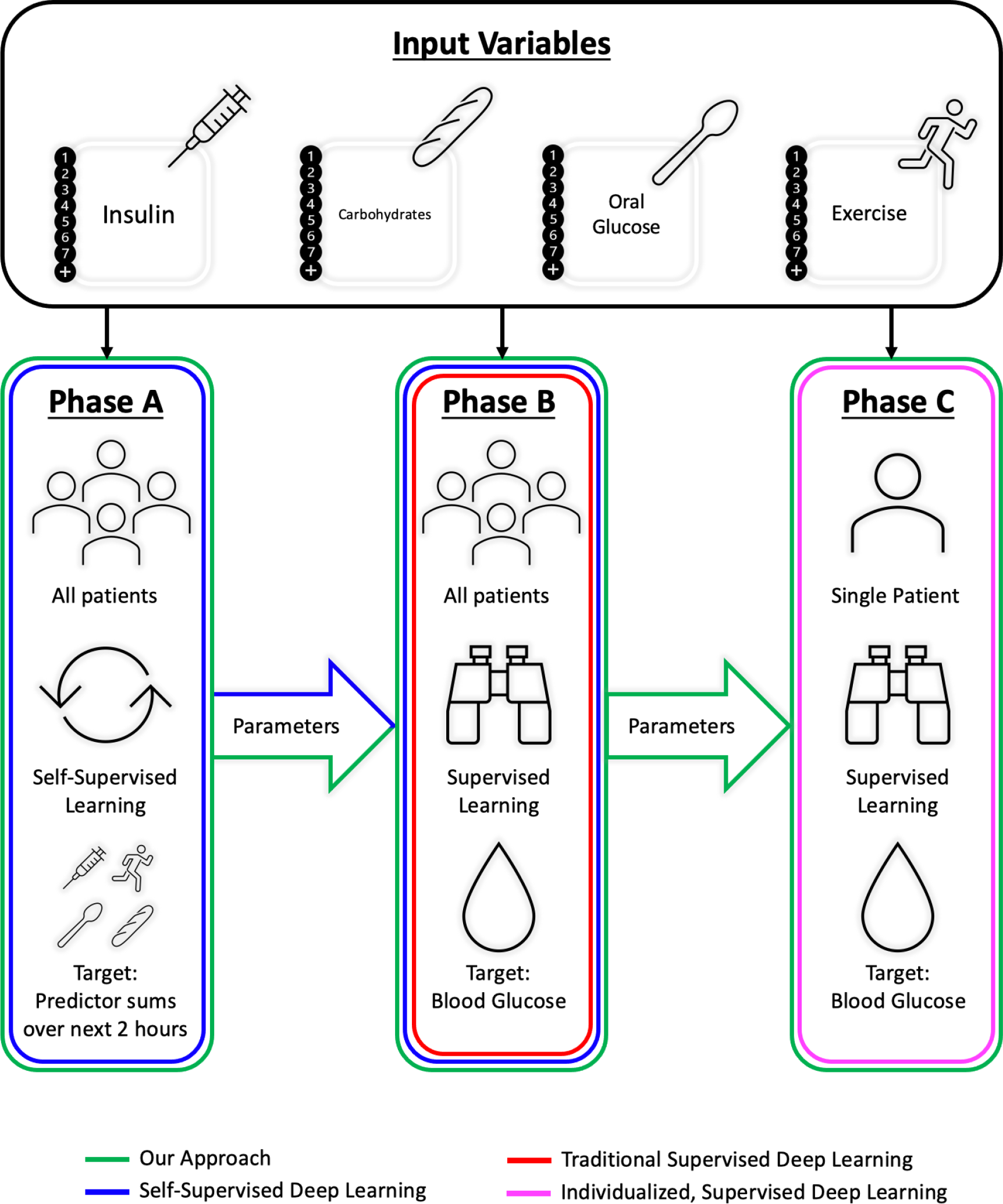
The Training Phases of our Novel Approach: First we perform a phase of self-supervised learning on the abundant predictor data of all patients. During this phase, the task is to predict the four sums of our predictors over the next 24 entries. This is followed by a phase of traditional supervised learning on all patient’s blood glucose labels. Finally, we leverage individualization with a final training phase that fine-tunes on the individual patient’s blood glucose labels. Other approaches tested are also highlighted.

An important aspect that separates the supervised training phases from the unsupervised phase in our approach is the loss function we employ. During self-supervised learning, we use mean-squared error (MSE), which is the default choice for many regression tasks. During supervised training, however, we use a custom loss function. Although blood glucose prediction, like our self-supervised task, is a regression problem, MSE is an inappropriate error metric here for medical reasons. Extreme blood glucose levels correspond to hypo- and hyperglycemia, conditions diabetics must avoid at all costs [39]. As such, an error of 20 in the 50 mg/dL range is far more costly than an error of the same magnitude in a healthier blood glucose range of around 150 mg/dL. MSE, however, would assign both cases the same loss, which is undesirable. To avoid this issue, we leverage a custom loss function called glucose-specific MSE (gMSE) developed by Del Favero, Facchinetti, and Cobelli (2012) [40], which disproportionately penalizes prediction errors in medically dangerous areas. The formula for gMSE is 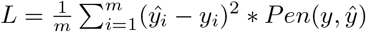, where Pen is a complex penalty term that depends on the medical danger associated with the model prediction and true label. The penalty function is defined as follows:

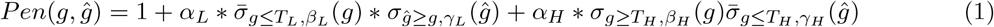

The intuition behind this function is that discrete penalties (*alpha*_*L*_ and *alpha*_*H*_) apply to certain areas that are particularly unhealthy to the patient. This allows a differentiation between false positive and false negative hypo-, and hyperglycemia-predictions. The bounds are defined by *T*_*L*_ and *T*_*H*_. The additional parameters define the sigmoid-approximations, which are introduced to ensure differentiability, which would not be the case for simple two-dimensional step-function.

The sigmoid approximations of the gMSE loss function are defined as follows:

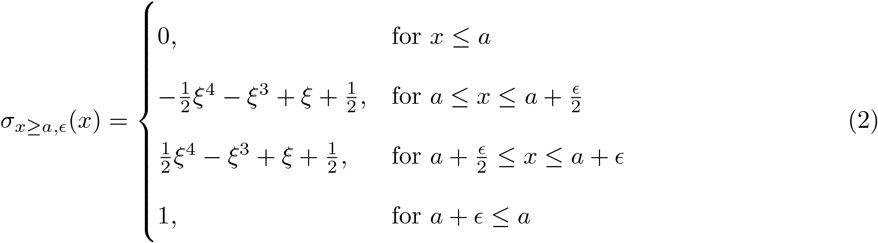

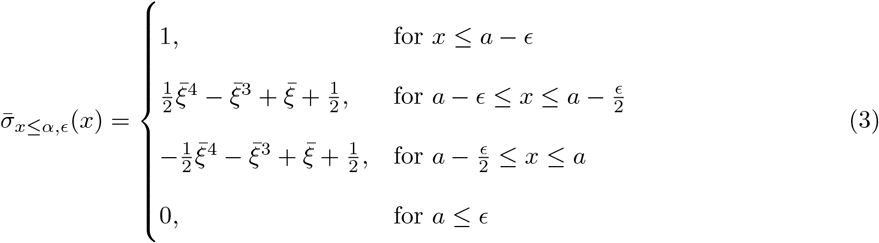

where *ξ* and 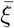 are defined as

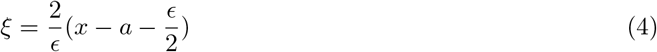

and

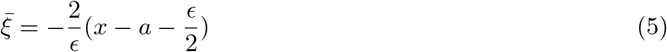

### Baselines

In order to evaluate and compare the performance of our novel training approach, we train and run identical experiments using four other training approaches. The approaches differ with respect to the presence of individualization and self-supervised learning, allowing for clear comparisons to ascertain the effectiveness of each facet of the training. By altering between adding a self-supervised learning stage and individualizing our model, we experiment on four distinct baseline training approaches. For all baseline approaches, as well as our final approach, we hold the model architecture constant.

The first baseline we compare to is traditional supervised deep learning. This is the simplest form of deep learning. We simply use the predictor data for all patients to train the model using gMSE in a single training phase and use the results to predict the respective blood glucose levels in the data. This approach involves neither self-supervised learning, nor individualization.

The second baseline we compare to is self-supervised deep learning. This is identical to traditional, supervised deep learning (baseline 1), except that we introduce a phase of self-supervised learning before the phase of traditional supervised learning. This baseline approach is very similar to our proposed new approach, with the exception that it makes no use of individualization. It uses all patients’ data, as well as the same model for predictions on all patients. While our approach also uses all patients’ data, its third training phase means that it yields a distinct model per patient, which is not the case here.

The third baseline we compare to is individualized, supervised deep learning. This is similar to traditional, supervised deep learning, with the sole difference being that only the data points of one patient are considered for predictions on that patient. Thus, this approach trains a different model for every patient based on completely distincts datasets, whilst traditional deep learning uses the same model for all patients. This baseline includes no self-supervised learning.

The fourth baseline we compare to is fully individualized, self-supervised deep learning. This is similar to individualized, supervised deep learning, with the addition of an individualized self-supervised training phase before the individualized supervised training phase. This baseline results in a distinct model for every patient trained on only its respective patient’s data. This model makes no consideration of the trade-off between individualization and self-supervised learning discussed earlier.

### Experiments

To evaluate the performance of our proposed approach, we run the same experiments for all approaches outlined in the previous sections. In each experiment, we first train the model on the first 80% of the dataset and use the resulting model to test on the remaining 20%. For individualized models, the predictions are based on the data of the respective individual patients, meaning we create 14 different models as part of the each experiment and aggregate their predictions for evaluation.

In order to assess the performances of our five model types, we run the above training and testing regime under varying levels of data availability, as this is a key aspect of our use-case. Rather than measuring blood glucose every 5 minutes as was done in the study we source our data from, we envision a scenario where that necessity is reduced to hourly, daily or weekly measurements. In order to simulate such an environment, we test the performance of our approaches by gradually reducing the availability of labels in the training dataset. We do this by keeping only every n^th^ row of our training glucose labels in the respective run of our experiment. We begin our experiments by keeping only every tenth label, which, since glucose measurements are around 5 minutes apart, apptoximately reflects a single glucose measurement every hour, and continue to drop labels throughout experiment runs until we keep only every 2000^th^ label, which translates to approximately one glucose measurement per week. Across all training approaches, this results in 50 experiments. For each experiment we record the gMSE, as well as the rMSE, and all individual predictions.

Throughout all experiments, we use hyperparameters chosen by training on the first 80% of the training dataset and evaluating performance on the last 20% of the training dataset. We use the sigopt library for tuning. For each of the baseline approaches, as well as our final approach, we tune different learning rates, and epoch numbers for every training stage, which reflects their varying natures. We also train a different dropout probability for every approach.

## Results

The results of our experiments are summarized in Table 1. Out of the evaluated training approaches, our approach was consistently the best performing, followed by standard self-supervised deep learning, which includes self-supervised deep learning, but not the added individualization stage our approach uses. A graphical representation of these results can be found in Figure 3. We also show the rMSE values in Figure 6 for interpretability. Here, self-supervised deep learning is more similar in performance to our final approach, which shows that our final training approach allowed models to better adapt to the custom loss function. Figure 5 provides a detailed scatterplot example of the predictions when only every 2000th row for supervised learning is available, which corresponds to roughly one blood glucose measurement per week.

**Table 1.** Performance of tested training approaches (gMSE)

| Data Missingness | 10 | 20 | 50 | 100 | 200 | 400 | 800 | 1000 | 2000 |
| --- | --- | --- | --- | --- | --- | --- | --- | --- | --- |
| Approx. Label Frequency | 50 min | 1h 40min | 4h 10min | 8h20min | 16h 40min | 1.375 days | 2.75 days | 3.5 days | 7 days |
| Sup. DL | **4016.50** | 5087.08 | 4844.43 | 4915.19 | 5294.20 | 5725.32 | 6088.29 | 10744.22 | 8191.23 |
| Self-sup. DL | 6646.09 | 6975.04 | 6925.94 | 5248.57 | 5977.61 | 6293.60 | 6805.33 | **5236.19** | 8206.98 |
| Indiv., sup. DL | 7002.01 | 6567.38 | 6998.47 | 9779.41 | 11050.86 | 10557.30 | 8392.47 | 10314.03 | 10739.87 |
| Fully indiv. self-sup. DL | 6513.66 | 6575.21 | 9298.39 | 6597.57 | 6692.13 | 12366.85 | 14536.41 | 20663.22 | 23313.15 |
| Our approach | 4685.12 | **4601.09** | **4547.19** | **4174.30** | **4502.65** | **4734.03** | **5139.91** | 5686.26 | **6387.47** |

For each approach we test performance under increasingly sparse data, keeping only the $n^{\text{th}}$ blood glucose label of the training data (data missingness). Best performance highlighted in bold. Self-supervised deep learning or our approach performed best in all experiments, with our approach particularly strong when data is very scarce.

**Fig 3.**
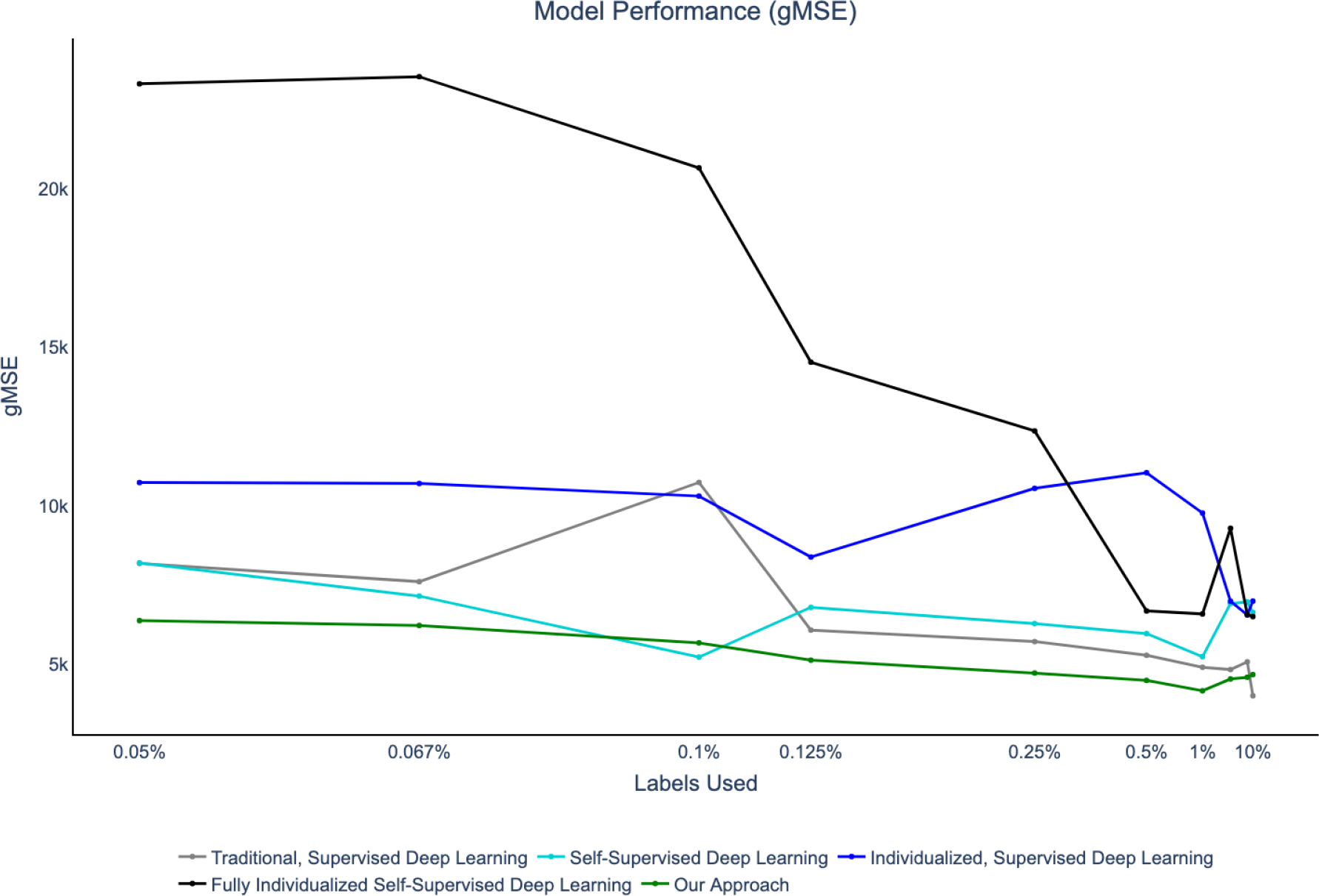
Model Performance under different training approaches in gMSE: Test gMSE of baseline approaches and our approach for different levels of data label availability. (see Table 1).

**Fig 4.**
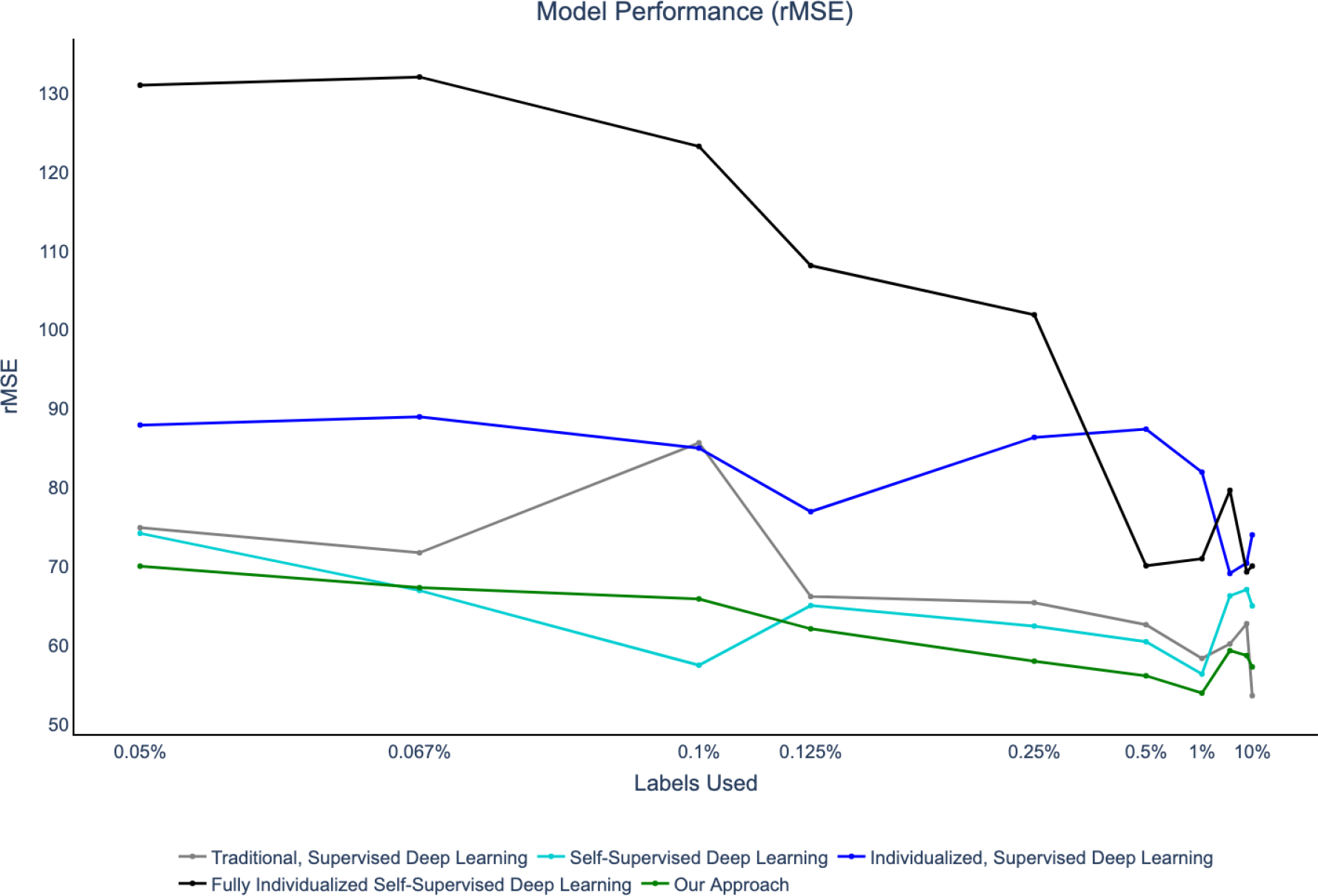
Model Performance under different training approaches in rMSE: Test rMSE of baseline approaches and our approach for different levels of data label availability. Included for interpretability.

**Fig 5.**
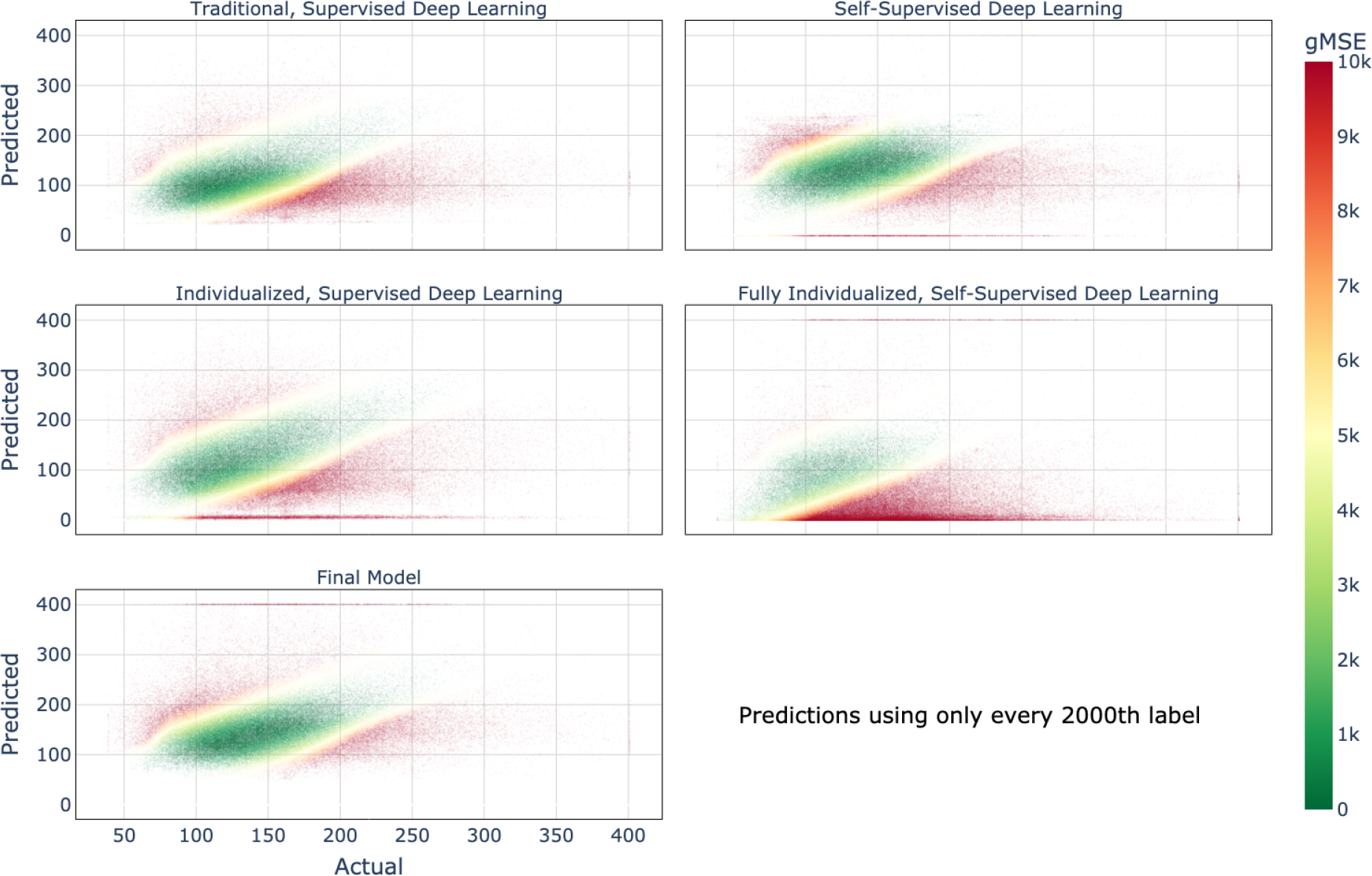
Predicted Glucose vs Actual Glucose when only every 2000th label is available: Color shading shows gMSE value. Keeping only every 2000^th^ label equates to roughly keeping only one blood glucose measurement per week.

**Fig 6.**
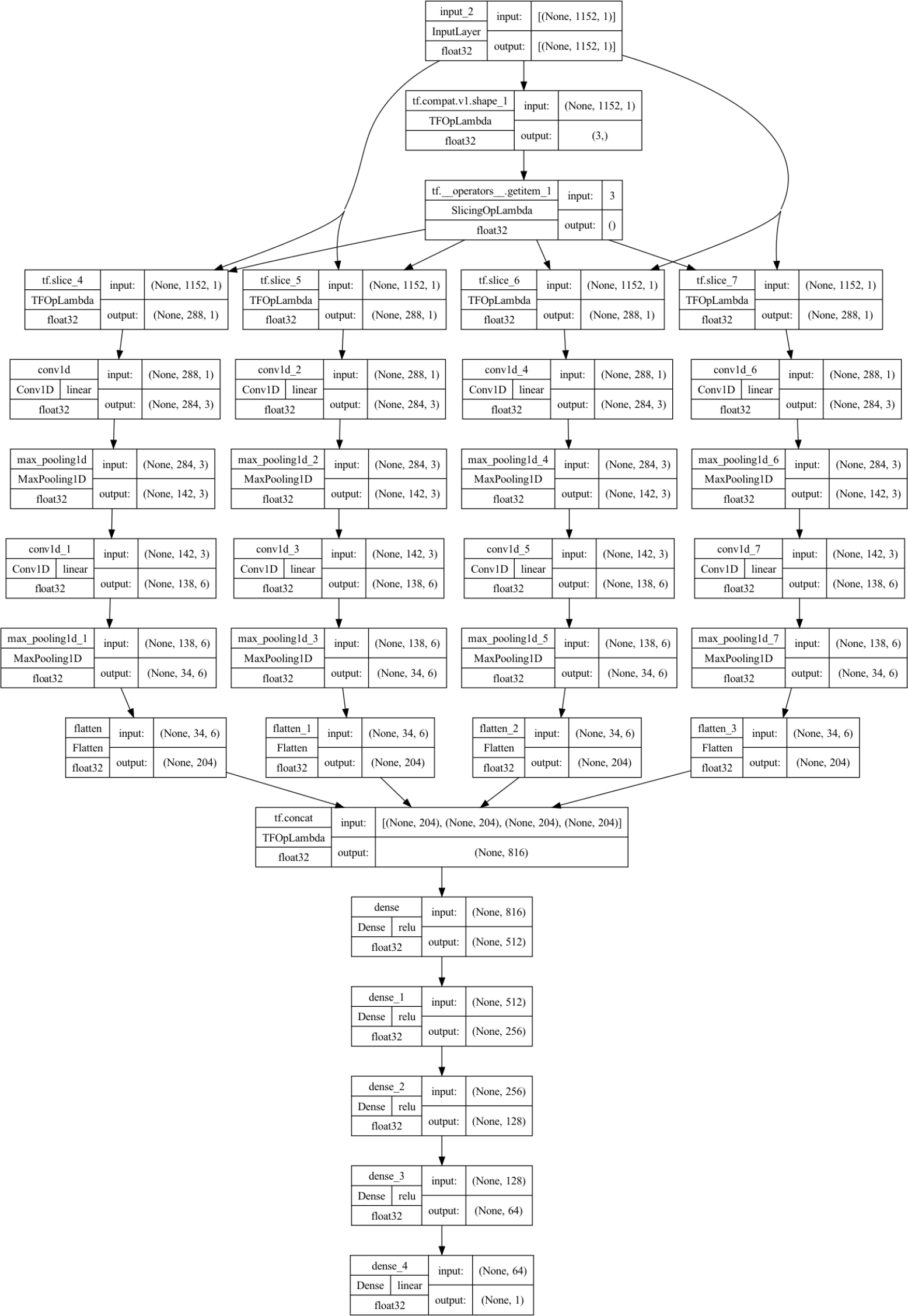
Fully detailed model architecture including layer sizes. Included for completeness.

## Discussion

We find that the gMSE performance on the test set generally decreases for all training approaches as we reduce the amount of training data. This effect and the differences between models become especially pronounced for models trained on very sparse data with at most 1% of the original blood glucose labels. This is also the regime that is the main focus of this study in order to minimize the need for invasive measurements. The results also demonstrate the overall effectiveness of combining self-supervised learning with individualization in blood glucose prediction.

### Self-Supervised learning

The comparison between Traditional Deep Learning (Baseline 1) and Self-Supervised Deep Learning (Baseline 2) clearly indicates the benefit that self-supervised learning across patients has in conjunction with supervised learning on a sparse dataset. With blood glucose labels available at a rate of 1-2 measurements per week, the gMSE is reduced from around 9000 to 6000, which is a substantial improvement, especially considering the limited scope of the predictor data we have at our disposal to perform self-supervised learning. The fact that fully individualized self-supervised deep learning performed so poorly, despite the presence of self-supervised learning on the far less abundant personal data, is likely a consequence of the fact that self-supervised learning requires a larger data volume to work well. A different interpretation of the poor performance here would be that in this use-case self-supervised learning may be uncovering universal features, rather than patient-specific idiosyncrasies, although this hypothesis would have to be tested against the case with similar data amounts at an individual level. In a study with richer predictor data, which could come from wearable devices for example, the potential for self-supervised learning to contribute to performance improvements is likely even greater than what we demonstrate here.

### Individualization

Training the model on observations from only one patient throughout all stages does not yield performance comparable to training a model on a large body of non-individualized data, as the sub-par performances of our individualized baselines show. Despite this, the performance of our approach still demonstrates, that, similarly to how many modern large language models are finetuned to specific tasks, small adjustments based on individualized labels, can help to enhance downstream performance. In using individualization only for fine-tuning, and using all data for self-supervised learning, our training approach combines the most useful aspects of individualization and self-supervised learning. This results in the best performance of the entire study, confirming that both major ideas of this study can jointly lead to better prediction performance and therefore improved patient health in settings with a limited availability of invasive data.

### Limitations

This study is only intended to demonstrate the potential of the self-supervised learning and individualization for moving towards clinically usable accuracy of blood glucose levels. Given the simple nature of the four predictors we use as well as the low number of patients, a major limitation of this study is therefore that its results are not clinically usable. A further limitation of this study is that even though we use only non-invasive predictors, we still rely on a low number of blood glucose samples to make our approach work. So long as blood glucose sampling remains an invasive exercise, this approach will not be entirely non-invasive by extension. Furthermore, the results shown in this study assume a high level accuracy of the CGM devices that were used to collect the blood glucose data, which may not be present. Flawed measurements, or a variance in the measurements, will translate to worsened predictions of the trained model. Another limitation of this study is that due to computational constraints, we were not able to repeat our calculations with different random seeds to calculate confidence intervals of our findings. We therefore need to rely on our results as point estimates for the ground truth improvement of self-supervised individualized learning over traditional deep-learning approaches.

### Future Work

Further research into the potential for the replacement of CGM devices through deep learning can build on this study in various ways. First, more data about nutrition and vital signs, such as heart rate or body temperature could be used as predictors. Datasets from smart devices, such as personal fitness watches, may offer a promising avenue for this in the future, and recent preliminary works support this potential [42–44]. Secondly, new datasets may offer potential for more well-suited tasks in self-supervised-learning. The task we use is an artifact of the dataset at hand, and based on the intuition that the sum of predictors over the next two hours may encode things such as biorhythm or daily routines. In other datasets with more predictors, there may be tasks that contain more relevant information for downstream learning and whose basis lies in biological arguments rather than high-level intuition. Finally, there are other architectures we could have chosen for this study. It may be the case that other model architectures respond more favourably to self-supervised learning than ours. RNNs, LSTMs, or transformers could all prove to be superior alternatives and should be investigated.

## Data Availability

All data files are available for download at https://public.jaeb.org/jdrfapp2/stdy/465

## Acknowledgments

The authors would like to thank Kari Hanson for organizing the Stanford ICME XPlore program. This program has played a tremendous role in supporting this research and has provided valuable access to the ICME cloud computing cluster.

